# Uncertainty-aware extraction of clinical findings from Finnish EHRs using open large language models

**DOI:** 10.64898/2026.07.07.26355248

**Authors:** Jussi Leinonen, Juha Knuuttila, Siina Pamilo, Samu Kurki, Miika Koskinen

## Abstract

**Objective:** To evaluate whether open-weight large language models (LLMs) can accurately extract clinical findings from Finnish-language pediatric records, and whether prediction uncertainty can be used to triage cases for expert review to minimize manual work

**Materials and Methods:** Retrospective cohort of 97 pediatric ischaemic stroke patients (1 month–17 years) from Helsinki University Hospital (2010–2023). Three open LLMs (gpt-oss-20b, DeepSeek-R1-Distill-Qwen-32B, and medgemma-27b-text-it) were prompted in English to detect four extraction targets (hemiplegia, headache, seizure, and stroke as a positive control) from each patient’s full free-text record. Each combination received 15 calls (five temperatures × three repeats). Performance was benchmarked against a clinician reference (accuracy, recall, precision, F1). Shannon entropy across the 15 calls quantified within-model uncertainty; inter-model disagreement provided an ensemble signal. Patients were ranked by uncertainty for a simulated selective-review workflow. Findings were externally validated in an independent neonatal stroke cohort (n = 88).

**Results:** Gpt-oss-20b achieved the best balance of recall (0.91–1.00) and precision (0.83–0.92), with F1 0.89–0.95 across non-control extraction targets. Entropy in misclassified cases was 2.4–3.4 times higher than in correctly classified cases. Entropy-based triage achieved complete error coverage by reviewing <10% of patients for hemiplegia (8.3%) and headache (8.2%), and 19.6% for seizure. Neonatal validation reached F1 0.95 for Apgar 1 min and binary seizure, and F1 0.87 for 4-class stroke-subtype classification.

**Discussion:** Within-model entropy and inter-model disagreement provided complementary, calibrated signals of likely error in a non-English clinical setting.

**Conclusion:** Open LLMs can extract clinical findings from Finnish pediatric records with accuracy comparable to published English benchmarks, and uncertainty-based triage substantially reduces required expert workload.

## 1 Introduction

Most clinical information is still recorded in free-form text, whether in specialist-care notes, pathology reports, or nursing records. This is particularly true of symptom descriptions, status documentation, and clinical reasoning, which are seldom captured as structured codes or tables. As a result, systematic patient-level use of this information for research and quality monitoring still relies on labour-intensive manual extraction, even though electronic health records (EHRs) have resolved some of the paper-era challenges. Manual review of a single patient’s records can take as long as a clinic visit, which makes systematic analysis of large cohorts effectively infeasible without automation.

Recent developments in large language models (LLMs) have substantially changed this picture. Several studies have shown that LLM-based methods can match or exceed traditional approaches for extracting structured data from clinical text. Azar et al. [1], using a GPT-4–based NTEP tool, achieved 99.8% classification accuracy against a manually curated reference for variables extracted from radical-prostatectomy pathology reports; the model was deployed in a secure, closed environment. Dao et al. [2] developed an open Llama-based generative extraction method for right-heart catheterisation procedure notes and achieved 99% precision, 85% recall, and an F1 of 91.5% for haemodynamic variables across 200 procedure records, with low hallucination rates. Alkhalaf et al. [3] used an open Llama 2 model with retrieval-augmented generation (RAG) to analyse nutrition notes from Australian residential aged-care units; zero-shot prompting produced structured nutritional summaries at 93.25% accuracy and identified malnutrition-related factors at approximately 90% accuracy compared with assessments by dietitians and nurses.

Submitting an entire text document to an LLM is not necessarily the most effective way to use the models on long clinical records. Long documents may in some cases exceed the model’s context window, and performance can degrade well before the context limit is reached, when the model fails to use information distributed evenly throughout the document [4]. Lopez et al. [5] presented Clinical Entity Augmented Retrieval (CLEAR), in which LLM-assisted entity recognition, ontology-based expansion, and targeted retrieval are combined so that the model sees only the clinically relevant portions of the text. CLEAR achieved higher F1-score for clinical-variable extraction than conventional whole-document processing, while reducing token use and computation time by more than 70%. This supports the idea that entity-driven, selective context construction can improve both performance and efficiency for long clinical texts. However, contextual information may be lost in this process, and for that reason our analysis processed the entire patient record at once, accepting longer inference times in exchange for avoiding context loss.

Although the existing literature already covers several medical specialties and approaches, the behaviour of LLMs on non-English clinical data has been studied less extensively. Most published studies use English-language data, and validation on minority languages such as Finnish clinical text has not yet been reported. Furthermore, most work focuses on extraction accuracy and addresses less how a model’s intrinsic uncertainty, agreement between models, and text length affect reliability and the need for additional manual work. Recent work has begun to examine the ability of models to evaluate their own and other models’ output: Chung et al. [6] described the VeriFact system, in which a multi-model “LLM-as-a-Judge” approach achieved approximately 93% agreement with clinician judgements when fact-checking patient-level summaries. This highlights that LLMs can also be used to assess claims and reasoning produced by other models.

In this study we examined the ability of three open LLMs (gpt-oss-20b, DeepSeek-R1-Distill-Qwen-32B, and medgemma-27b-text-it) to identify four key extraction targets (hemiplegia, headache, seizure, and stroke) in a Finnish pediatric-stroke patient cohort. All study patients had a confirmed stroke diagnosis, which served as a positive control for the technical functionality of the models. Hemiplegia, headache, and seizure represented clinically and linguistically more variable extraction targets. Because LLM outputs are non-deterministic, the same model may answer differently when the same patient record is queried repeatedly. This behaviour resembles multiple clinicians reading the same notes and occasionally disagreeing. Thus, we analysed model uncertainty using entropy and prediction agreement across different runs and assessed how these uncertainty signals and multi-model ensembling can be used to focus manual review. Our work complements the prior literature in three ways: (1) by bringing the pediatric-stroke perspective to LLM-based information extraction, by introducing the use of multiple LLMs as a tool for uncertainty-based error identification, and by evaluating method performance on long-form Finnish-language free text.

## 2 Methods

### 2.1 Study population and data

This was a retrospective registry study using electronic health record (EHR) data from Helsinki University Hospital (HUS), collected between 1 January 2010 and 31 December 2023. Eligible patients were aged 1 month to 17 years (n = 97) with a primary ICD-10 diagnosis of cerebral infarction (I63), occlusion of cerebral arteries without infarction (I66), other cerebrovascular disease (I67), or sequelae of cerebrovascular disease (I69). Primary intracerebral and subarachnoid haemorrhages were excluded because their clinical presentation, aetiology, and symptomatology differ substantially from those of ischaemic stroke. HUS is a tertiary academic medical centre serving a catchment area of approximately 2.2 million inhabitants and more than one third of the Finnish paediatric population. Detailed clinical characteristics of this patient population, including aetiology, treatment, and outcomes, have been described separately in a companion population-based study [10].

Pseudonymised patient-level data were retrieved from the EHR system. Structured data included demographics, diagnoses, imaging studies, and treatments. Free-text paediatric, neurology, and paediatric-neurology notes were reviewed both to confirm the stroke diagnosis and to extract the clinical findings of interest. To improve epidemiological coverage, diagnosis ascertainment was supplemented by systematic review of MRI reports, which allowed identification of patients who lacked a specific cerebral-infarction diagnosis code. This step was intended to mitigate the under-ascertainment typical of retrospective code-based studies.

External validation was performed in a separately curated neonatal stroke cohort (n = 88), assembled using the same selection criteria as the primary cohort.

All analyses, including all LLM inference, were carried out within the secure HUS Academic computing environment. The study complied with the Finnish Act on the Secondary Use of Health and Social Data (552/2019) and was approved by HUS (HUS/265/2023).

### 2.2 Extraction targets and annotation

Four clinically relevant extraction targets were defined: hemiplegia, headache, seizure, and ischaemic stroke. As all study patients carried a stroke diagnosis, the stroke target served as a positive control, near-perfect performance was therefore expected on this target.

For each extraction target, an English-language clinical definition was drafted that specified which mentions in the text qualified as positive and which were excluded. For example, isolated sensory symptoms without motor deficit, or symptoms occurring in the distant past, did not satisfy the positive criteria for hemiplegia.

These definitions guided both the human reference annotation and the LLM prompt. Reference annotation was performed by a single clinical annotator within each cohort; the primary cohort and the validation cohort were annotated by different clinicians, providing partial inter-annotator perturbation across the validation boundary.

### 2.3 Language models and prompt design

We evaluated three open-weight LLMs (Table 1): OpenAI’s gpt-oss-20b [7], DeepSeek-R1-Distill-Qwen-32B [8], and Google’s medgemma-27b-text-it [9]. All three were openly available as of autumn 2025. Each model was required to be deployable within the secure HUS Academic environment on a single NVIDIA H100 GPU using Python.

**Table 1.** Language model characteristics.

| Model | Parameters<br>(total /<br>active) | Architecture | Type | Notable<br>features | Reference |
| --- | --- | --- | --- | --- | --- |
| gpt-oss-20b | 21 B / 3.6 B | MoE<br>Transformer | Open-weight | OpenAI’s<br>open-weight<br>model | [7] |
| DeepSeek-R1-Distill-Qwen-32B | 32 B | Transformer | Open-weight | Distilled<br>reasoning<br>model | [8] |
| medgemma-27b-text-it | 27 B | Transformer | Open-weight | Medical-<br>domain-<br>specialised<br>model | [9] |

For every patient, the model received the patient’s complete Finnish-language clinician notes and was asked to determine, for a given extraction target, whether that target was mentioned in the text. We used a single, task-uniform prompt template in English that placed the model in the role of a clinical assistant evaluating the target against pre-specified criteria, and that explicitly forbade inferences extending beyond the text. The output was constrained to a precise JSON schema containing a binary flag (1: identified, 0: not identified) and a brief English supporting summary. The supporting summary was used only as a qualitative aid for interpretation and did not enter the quantitative analysis. The prompt skeleton was identical across extraction targets; only the target name and its clinical description varied.

### 2.4 Performance metrics and prediction-stability assessment

#### Repetition design and temperature settings

The model performance was assessed for each of three models, four extraction-targets and five temperatures (0.5, 0.75, 1.0, 1.25, and 1.5), yielding 60 binary responses per patient. This design was used to enable assessment of prediction stability and uncertainty-based prioritisation of expert review.

Model output was evaluated along two axes: (i) classification accuracy relative to the clinician reference, and (ii) prediction consistency across repeated queries on the same case under varying parameter settings.

Each prediction event was defined by five factors: (i) language model, (ii) temperature, (iii) execution environment, (iv) prompt, and (v) input data (one patient’s free-text notes).

Classification performance was summarised with four standard metrics:

- **Accuracy**: overall fraction of correct predictions.
- **Recall** (sensitivity): ratio between predicted positive and true positive cases.
- **Precision** (positive predictive value): ratio between correctly predicted positive cases and all positive predictions.
- **F1**: a harmonic mean of recall and precision, summarising classification performance in a single value.

Because LLM outputs are non-deterministic, the same model can return different responses to the same patient on different repetitions. We assessed prediction consistency at two levels:

- *Within-model variation*. Each model was queried 15 times per patient and extraction target, as described above (three repeats at each of five temperatures), producing a distribution of binary responses. The uncertainty of this distribution was quantified using Shannon entropy [11], an established measure of informational uncertainty in both LLM research [12] and inter-rater agreement [13]. In practical terms, a value of zero indicates that the model constantly returned the same answer, whereas high entropy indicates randomness in responses.
- *Cross-model disagreement*. Predictions from different models were compared by computing how often pairs of models reached opposing answers for the same patient and extraction target. Each model’s final prediction was determined by majority vote across all 15 repetitions, mirroring the practice of summarising several independent assessments into a single most-likely answer.

### 2.5 Uncertainty-based targeting of expert review

Expert review of every case is rarely feasible in clinical text mining. We therefore evaluated whether model-derived uncertainty can be used to identify error-prone cases and to direct limited expert resources where they yield the greatest added value.

For each classification objective, a confusion matrix (true positives, false positives, true negatives, and false negatives) was constructed across patients. Then, within each combination of model, extraction target, and confusion class, patients were ordered by uncertainty, and consecutive patients were packed into bins. Each bin contained between three and five patients: groups of three to five patients formed a single bin, and larger groups were split into bins of predominantly three patients. No bin contained fewer than three or more than five patients, in compliance with the anonymization rules of the data. For each patient bin we computed two uncertainty measures:

- **Within-model uncertainty** — the consistency with which a single model produces the same prediction across repeated queries with varying parameters; high entropy indicates a labile prediction. Gpt-oss-20b was used as the base model for this analysis.
- **Inter-model disagreement** — how often the different models reach different answers for the same patient and extraction target, analogous to receiving conflicting opinions from multiple expert consultants.

We simulated selective expert review as follows. For each extraction target, patient bins were ranked from highest to lowest uncertainty. We then selected the most uncertain bins first and continued down the list until the cumulative number of patients reached each target review proportion (10%, 20%, or 30% of the cohort). Because bins contain 3–5 patients, the realised review proportion could fall slightly above or below the nominal target. If uncertainty is informative, misclassifications should concentrate in the highest-uncertainty bins. A strategy is efficient when a small, reviewed fraction captures a large share of errors.

## 3 Results

### 3.1 Text length and number of records

Records from 97 pediatric stroke patients were analysed, constituting a population-based sample from Southern Finland with an incidence of approximately 2.5 in 100 000 children. All free-text notes for a given patient were concatenated into a single document. The length of the analysed patient texts varied substantially. Most texts contained between 10,000 and 20,000 characters, and approximately one third were longer than this (Figure 1). One A4 page of Finnish free text corresponds to roughly 2,000–2,500 characters, corresponding to 4–10 pages. This text length requires the model to handle long, clinically multidimensional documents.

**Figure 1.**
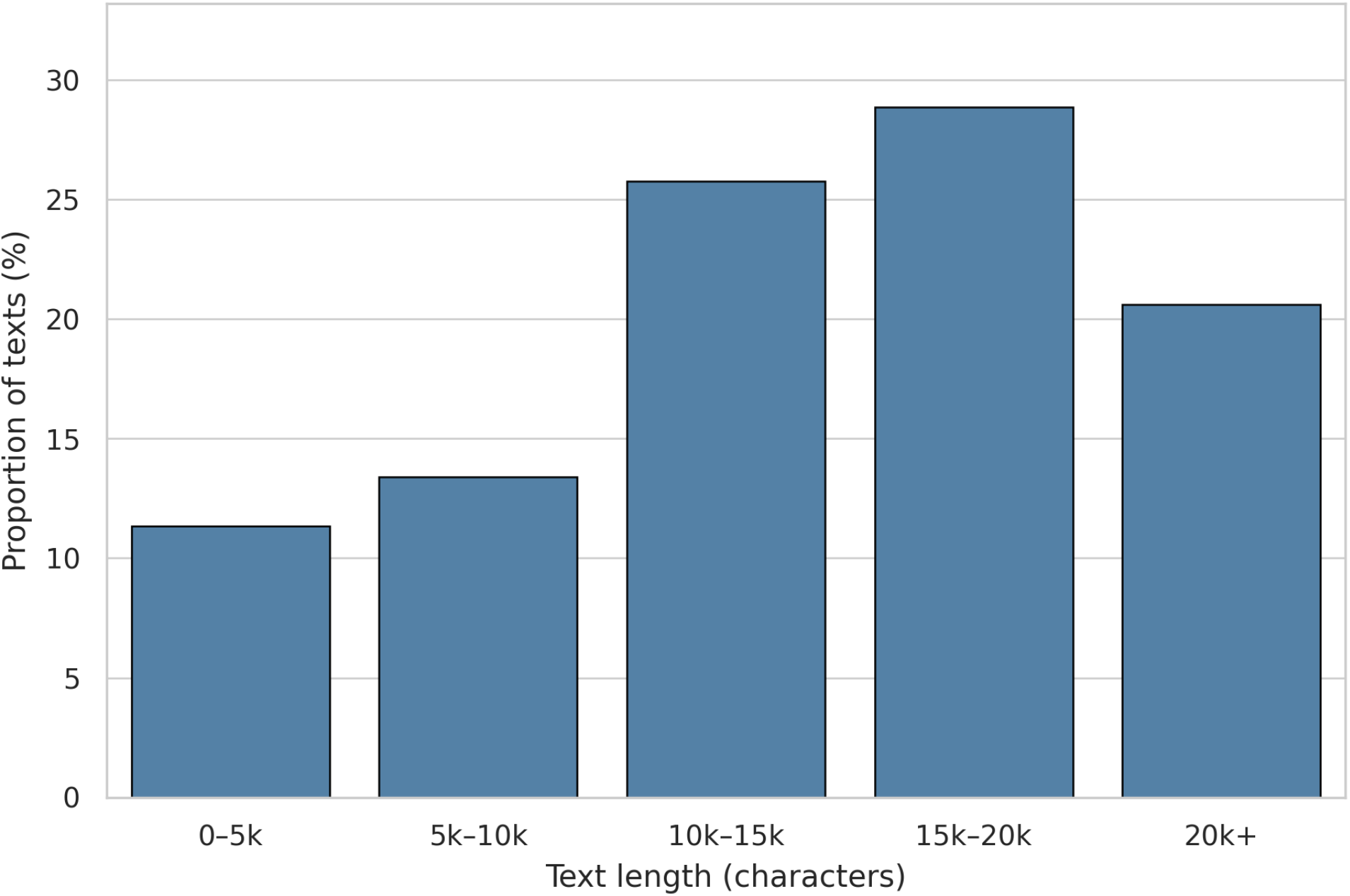
Length distribution of the analysed patient texts. Distribution of patient-level text length, in characters, across binned length classes for the n = 97 primary-cohort patients. All free-text notes for a given patient were concatenated into a single document before counting. As a rough conversion, one A4 page of Finnish free text corresponds to approximately 2,000–2,500 characters.

### 3.2 Classification performance

Table 2 summarises the models’ performance metrics by extraction target. Gpt-oss-20b achieved the highest F1-score across more challenging extraction targets: headache (0.91), hemiplegia (0.95), and seizure (0.89). The stroke target served as a positive control, on which all models achieved high performance (F1 0.95–0.98).

**Table 2.** Model performance metrics and entropy (uncertainty) by extraction target. F1 = F1 score; Recall = sensitivity; PPV = positive predictive value (precision); Accuracy = overall classification accuracy; Entropy = model-level patient-weighted mean Shannon entropy across the 15 calls per patient x extraction target, computed separately for each model as the patient-count–weighted mean of per-patient entropies across the cohort (weights = number of patients in each 3–5-patient analysis bin).

| Extraction target | Metric | DeepSeek-R1-Distill-Qwen-32B | gpt-oss-20b | medgemma-27b-text-it |
| --- | --- | --- | --- | --- |
| Headache | F1 | 0.84 | **0.91** | 0.81 |
|  | Recall | 0.84 | **1.00** | **1.00** |
|  | PPV | **0.84** | 0.83 | 0.68 |
|  | Accuracy | 0.92 | **0.95** | 0.88 |
|  | Entropy | 0.167 | **0.050** | 0.288 |
| Hemiplegia | F1 | 0.86 | **0.95** | 0.85 |
|  | Recall | 0.95 | **0.98** | **0.98** |
|  | PPV | 0.79 | **0.92** | 0.75 |
|  | Accuracy | 0.82 | **0.94** | 0.80 |
|  | Entropy | 0.305 | **0.137** | 0.246 |
| Seizure | F1 | 0.60 | **0.89** | 0.76 |
|  | Recall | 0.77 | 0.91 | **0.95** |
|  | PPV | 0.49 | **0.87** | 0.64 |
|  | Accuracy | 0.76 | **0.95** | 0.87 |
|  | Entropy | 0.421 | **0.098** | 0.192 |
| Stroke | F1 | **0.98** | 0.95 | **0.98** |
|  | Recall | **0.97** | 0.91 | 0.96 |
|  | PPV | **1.00** | **1.00** | **1.00** |
|  | Accuracy | **0.97** | 0.91 | 0.96 |
|  | Entropy | 0.101 | 0.113 | **0.077** |

The clearest difference among models was the recall–precision trade-off. Medgemma-27b-text-it had high recall (0.95–1.00) but lower PPV on the non-control targets (0.64–0.75), whereas gpt-oss-20b combined high recall (0.91–1.00) with the best PPV (0.83–0.92) and the highest accuracy on headache, hemiplegia, and seizure. DeepSeek-R1-Distill-Qwen-32B lagged on seizure (F1 0.60; PPV 0.49) but tied Medgemma on the stroke control (F1 0.98).

### 3.3 Prediction uncertainty

Table 2 also reports patient-weighted entropy by model and extraction target, describing each model’s intrinsic prediction stability: the smaller the value, the more consistently the model produces the same answer across repeated calls. Gpt-oss-20b had systematically the lowest entropy on the more challenging extraction targets (headache 0.05; hemiplegia 0.14; seizure 0.10). DeepSeek-R1-Distill-Qwen-32B was the most uncertain, particularly on seizure (0.42); medgemma-27b-text-it was intermediate. On the stroke positive control, all models had low entropy (0.08–0.12), consistent with the expected high prediction certainty.

Entropy in misclassified cases was substantially higher than in correctly classified cases for all extraction targets (Table 3). For hemiplegia, the mean entropy of misclassified cases (0.53) was nearly threefold that of correctly classified cases (0.19). The same pattern was observed for headache (0.46 vs. 0.13), seizure (0.49 vs. 0.20), and stroke (0.31 vs. 0.09). These results indicate that model prediction uncertainty is an informative signal for identifying error-prone cases.

**Table 3.**
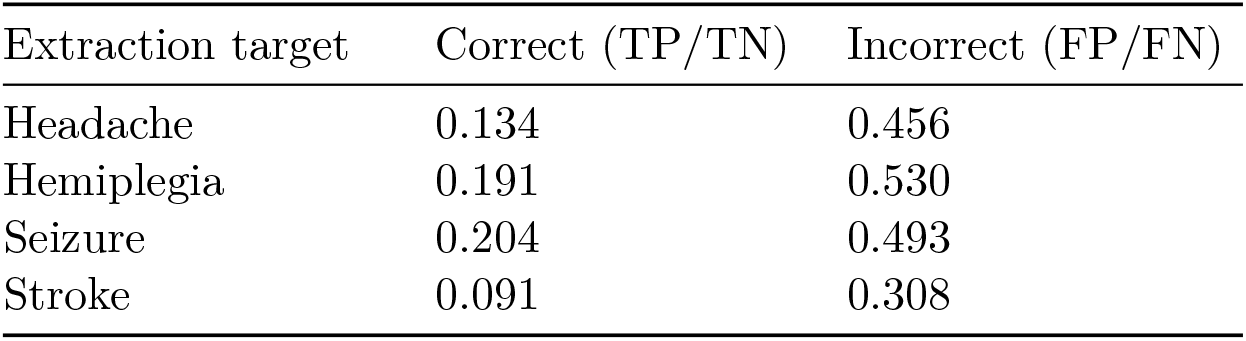
Entropy (uncertainty) in correctly (TP/TN) and incorrectly (FP/FN) classified cases. For each extraction target, cases were split into correct (TP/TN) and incorrect (FP/FN) using gptoss-20b’s majority vote against the clinician reference. Within each split, a patient-count–weighted mean entropy was computed separately for each model (weights = patients per 3–5-patient analysis bin); the table reports the unweighted arithmetic mean of those three model-level values. FN = false negative; FP = false positive; TN = true negative; TP = true positive; NA = no cases in that class.

| Extraction target | Correct (TP/TN) | Incorrect (FP/FN) |
| --- | --- | --- |
| Headache | 0.134 | 0.456 |
| Hemiplegia | 0.191 | 0.530 |
| Seizure | 0.204 | 0.493 |
| Stroke | 0.091 | 0.308 |

### 3.4 Inter-model disagreement

Inter-model disagreement varied markedly (Table 4). Among true-positive (TP) classifications, disagreement was low (0.18–0.30), whereas among false-negative (FN) cases disagreement was almost universal (0.95–1.00). In other words, when a model failed to detect a true-positive case, the other models almost always disagreed — that is, at least one of the other models correctly identified the case. Similarly, false-positive (FP) cases showed high inter-model disagreement (0.65–0.94), reflecting divergent interpretations of borderline cases.

**Table 4.** Inter-model disagreement (ensemble) by extraction target and confusion-matrix class. For each extraction target and confusion-matrix class (FN, FP, TN, TP), each cell was assigned using gpt-oss-20b’s majority vote against the clinician reference. Values are the patient-count–weighted mean inter-model disagreement within that class (weights = patients per 3–5-patient analysis bin), where disagreement is the proportion of model pairs whose majority answers differ.

| Extraction target | FN | FP | TN | TP |
| --- | --- | --- | --- | --- |
| Headache | 0.950 | 0.652 | 0.370 | 0.251 |
| Hemiplegia | 1.000 | 0.869 | 0.547 | 0.194 |
| Seizure | 1.000 | 0.936 | 0.488 | 0.297 |
| Stroke | 0.975 | NA | NA | 0.184 |

*Note:* FP and TN are not applicable for the stroke positive control because all study patients had a confirmed stroke diagnosis (no true-negative cases by construction).

### 3.5 Targeting expert review

Figure 2 shows the minimum manual review required to catch all misclassified cases using uncertainty-based prioritization. Two different strategies were compared: within-model uncertainty (entropy of gpt-oss-20b across the 15 calls per patient × extraction target) and inter-model disagreement (ensemble disagreement across the three models). Each strategy was tested under three target review proportions (10%, 20%, and 30%); detailed results are given in Table 5.

**Table 5.** Targeting of expert review by extraction target and strategy. For each target review proportion (10%, 20%, 30%), the realised manual-review proportion and the cumulative coverage of misclassified cases (FP+FN) are reported. Man-% = realised manual-review proportion; Coverage-% = cumulative coverage of misclassified cases. Realised proportions can fall below or above the target because patients are processed in bins of 3–5.

| Extraction target | FP+FN (n) | Strategy | Target 10% |  | Target 20% |  | Target 30% |  |
| --- | --- | --- | --- | --- | --- | --- | --- | --- |
|  |  |  | Man-% | Coverage-% | Man-% | Coverage-% | Man-% | Coverage-% |
| Headache | 5 | Entropy | 8.2 | **100.0** | 17.5 | **100.0** | 29.9 | 100.0 |
| Headache | 5 | Ensemble | 7.2 | 0.0 | 19.6 | 0.0 | 27.8 | 100.0 |
| Hemiplegia | 5 | Entropy | 8.3 | **100.0** | 17.7 | **100.0** | 27.1 | 100.0 |
| Hemiplegia | 5 | Ensemble | 9.4 | 0.0 | 15.6 | 0.0 | 27.1 | 100.0 |
| Seizure | 7 | Entropy | 6.2 | 42.9 | 19.6 | **100.0** | 28.9 | 100.0 |
| Seizure | 7 | Ensemble | 9.3 | 42.9 | 19.6 | **100.0** | 28.9 | 100.0 |
| Stroke | 8 | Entropy | 9.3 | 0.0 | 17.5 | 62.5 | 29.9 | 100.0 |
| Stroke | 8 | Ensemble | 8.2 | **100.0** | 17.5 | **100.0** | 29.9 | 100.0 |

*Bold values indicate the smallest target proportion at which a given strategy first achieves complete coverage of misclassified cases for the row’s extraction target.*

**Figure 2.**
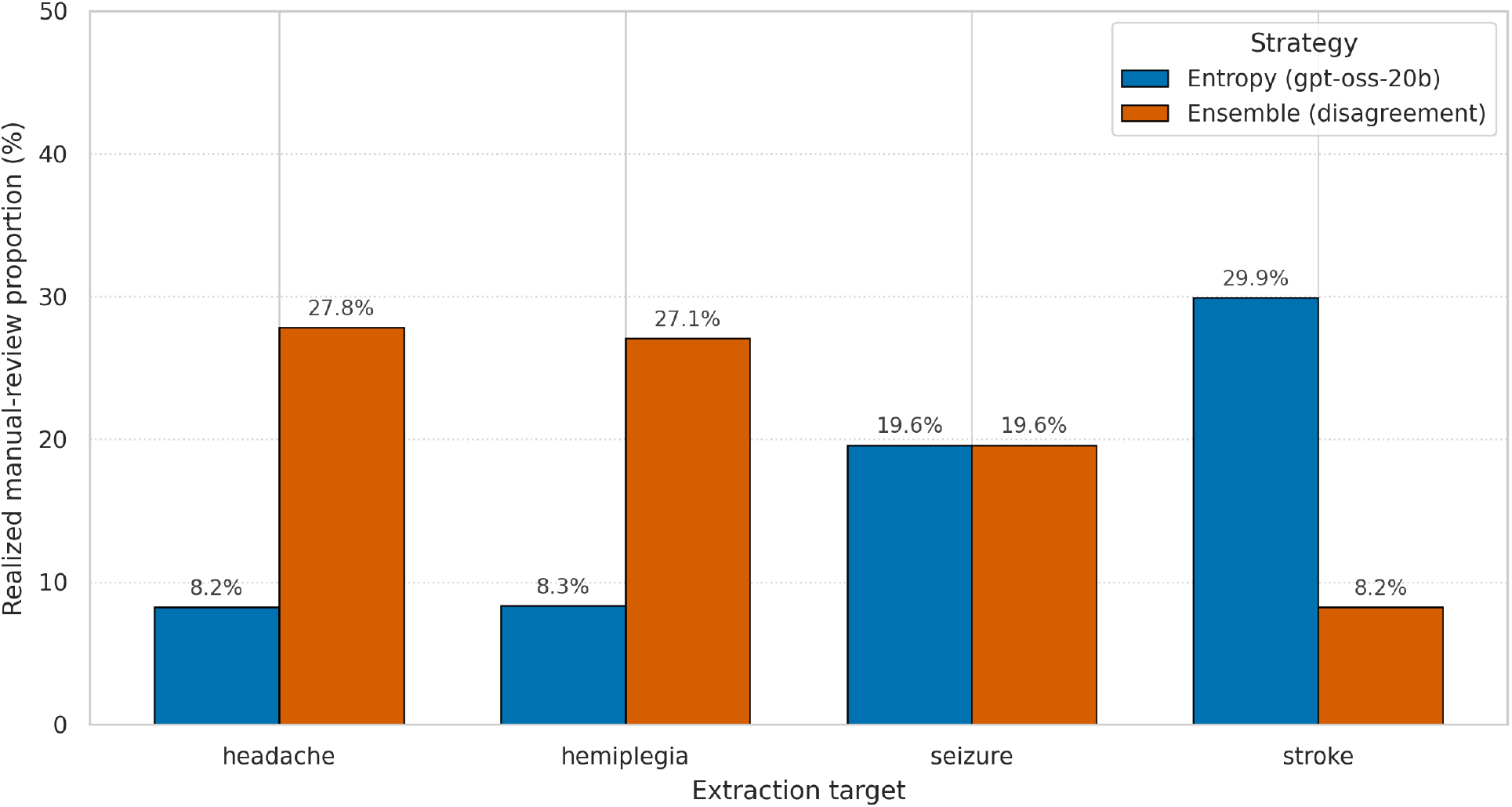
Targeting of expert review. The smallest manual-review proportion (%) needed to capture all misclassified cases (false positives plus false negatives) for each extraction target, under two prioritisation strategies. Blue bars: within-model uncertainty strategy (Shannon entropy of gpt-oss-20b across the 15 calls per patient × extraction target). Orange bars: inter-model disagreement strategy (ensemble disagreement across the three models). Detailed results at the 10%, 20%, and 30% target review proportions are reported in Table 5.

The within-model entropy strategy was particularly effective for headache and hemiplegia: already at the smallest target proportion (10%), all five misclassified cases were captured in the manually reviewed set, with the realised review proportion being only 8.2% (headache) and 8.3% (hemiplegia). For seizure, the 10% target captured 42.9% of errors (3/7 patients; the realised review proportion was 6.2%, falling below the target because patients are grouped in bins of 3–5), and complete error coverage was reached at the 20% target (realised 19.6%). On the stroke positive control, the entropy strategy was least efficient: at the 10% target, error coverage was 0%; at the 20% target, 62.5%; and complete coverage required the 30% target (realised 29.9%).

The inter-model disagreement strategy showed the opposite pattern across extraction targets. For stroke, complete error coverage was reached already at the 10% target (realised 8.2%). For seizure, the inter-model strategy performed identically to the within-model entropy strategy: 42.9% coverage at the 10% target and complete coverage at the 20% target (realised 19.6%). For headache and hemiplegia, however, the inter-model strategy directed no misclassified cases to manual review at the 10% or 20% targets (0% coverage), and complete coverage was reached only at the 30% target (realised 27.8% and 27.1%, respectively).

Overall, the within-model entropy strategy targeted manual review more efficiently for headache and hemiplegia (8% vs. 27–28%), whereas the inter-model disagreement strategy was clearly superior on the stroke positive control (8% vs. 30%). For seizure, both strategies performed equally well (20% target proportion).

### 3.6 External validation

Model performance was validated in the neonatal stroke cohort (n = 88) using three extraction targets (Table 6). The validation results show that the model can produce accurate structured information from clinical free text, particularly for binary and numeric extraction targets, whereas the multi-class stroke-subtype classification is clearly a more challenging task.

**Table 6.**
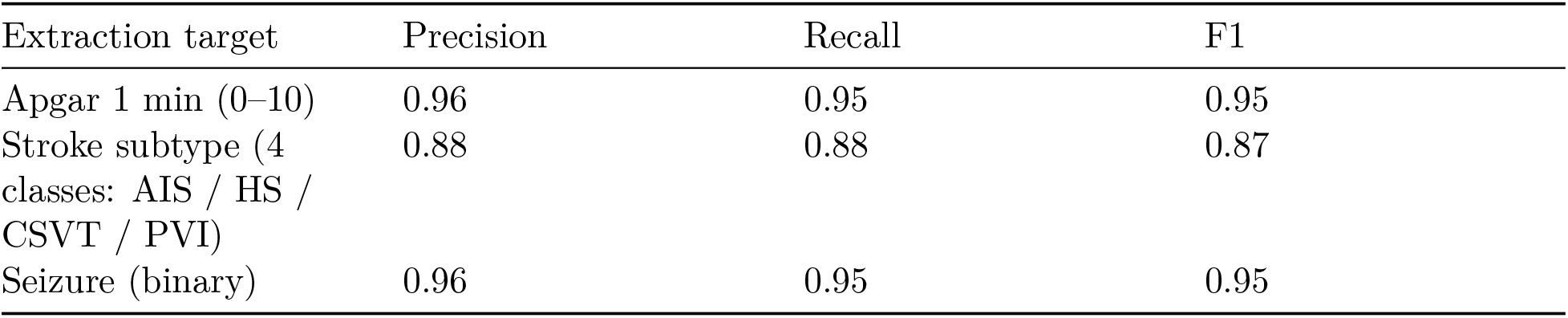
Model performance in the neonatal stroke cohort (n = 88) AIS = arterial ischemic stroke; HS = primary hemorrhagic stroke; CSVT = cerebral sinovenous thrombosis; PVI = periventricular venous infarction. The 4-class subtype label set comprises AIS, HS, CSVT, and PVI.

## 4 Discussion

### 4.1 Key findings

This work shows that LLM-based automatic recognition of clinical findings is feasible in a Finnish-language pediatric-stroke patient corpus. Of the three models evaluated, gpt-oss-20b offered the most balanced performance across all of the more challenging extraction targets, combining high recall (0.91–1.00) with the best precision (0.83–0.92; F1 0.89–0.95). It was also the most consistent in its predictions, as reflected in systematically the lowest entropy.

A key finding is that model prediction uncertainty — both within-model entropy and inter-model disagreement — correlates clearly with classification errors. The entropy of misclassified cases was several-fold that of correctly classified cases for every extraction target. This enables an uncertainty-based division of labour, in which expert review is targeted at the most uncertain cases. Our results show that already at less than 10% manual review, all hemiplegia and headache classification errors can be captured, and at approximately 20% review, all seizure classification errors as well, substantially reducing the total review burden.

### 4.2 Comparison with prior literature

Our findings are consistent with previous international studies in which GPT-4–based and open-LLM-based solutions have achieved high accuracy on a range of clinical information-extraction tasks, including pathology reports [1], right-heart catheterisation notes [2], and nutrition notes [3]. Our work extends this evidence by showing that comparable performance is achievable in a non-English, clinically specialised corpus.

In addition, the recognition capability of the models is not constant: it varies by extraction target. In the recent VeriFact study, Chung et al. [6] showed that LLM-based, patient-record-anchored fact-checking can achieve more than 93% agreement with clinical-consensus assessments and can produce more consistent evaluations than those of individual clinicians. This supports our observation that uncertainty signals and inter-model disagreement are not merely technical metrics but potential tools for prioritising clinical errors and quality-controlling automatically generated annotations.

### 4.3 Implementation and architectural choices

In this work, the entire written record of each patient was passed in a single LLM call, which in principle allows extraction of broader constructs than the simple presence or absence of an individual symptom or diagnosis. However, we restricted to the recognition of relatively simple concepts. In larger patient corpora, targeted extraction could be a more efficient alternative: the data could first be filtered with simple rule-based methods to text passages containing symptom-related keywords, with only those passages then passed to the LLM for analysis. This approach resembles the CLEAR procedure described by Lopez et al. [5], in which clinical entities serve as the basis for retrieval context and improve both performance and computational efficiency compared with whole-document processing.

We used moderately sized, pre-trained LLMs without task-specific fine-tuning. Substantially smaller models were also tried initially, but their performance was poor. This indicates that, in the absence of task-specific fine-tuning, model size has a clear impact on performance. On the other hand, the use of ever larger models increases compute time, memory, and energy requirements as well as inference time, all of which must be considered in a clinical production environment.

For these reasons it is unlikely that a practical solution would be to attach a single large LLM to every clinical use case. A more likely direction is a layered architecture in which a large model serves as a benchmark and possibly as a teacher for lighter models, but daily use relies on lighter, possibly smaller (distilled) or fine-tuned models combined with entity-driven retrieval context. Based on our results, a solution might emerge from a combination of lighter, locally deployable models, rule-based pre-filtering, and a well-designed uncertainty-based division of labour.

The task description (prompt) provided to the model was written in English even though the analysed data were in Finnish. Our results suggest that sufficiently large LLMs can process Finnish-language clinical text reliably with an English prompt. This makes international collaboration and similar analytics across language regions more accessible. It is nevertheless likely that models trained or fine-tuned on national clinical data could improve accuracy, particularly for extraction targets in which interpretation requires knowledge of Finnish clinical documentation conventions and terminology. The impact of prompt language is a planned ablation in future work.

### 4.4 Limitations

The study has several limitations. Although populationl represents approximately 1/3 of target population, the dataset was limited in size and originated from a single hospital organization (HUS), which limits the generalisability of the findings to other patient populations and documentation practices. The recognition of more complex clinical concepts — such as the temporal progression of symptoms, severity, or treatment response — may be substantially more challenging. The models were not compared with traditional rule-based or machine-learning text-mining methods, which would have offered an additional perspective on the relative benefit of LLMs. Reference annotation was performed by a single clinician within each cohort, although the two cohorts were annotated by different clinicians, providing partial inter-annotator perturbation across the validation boundary. The evaluation of uncertainty-based prioritisation was based on simulation, and its real impact on clinical workload and patient safety has not yet been assessed.

### 4.5 Future research directions

Future work should evaluate LLM-based concept recognition in larger, multi-centre datasets and on clinically more complex extraction targets. Of particular interest would be a comparison between the performance of pre-trained general-purpose models, models fine-tuned on Finnish clinical data, and traditional text-mining approaches. The benefit of uncertainty-based targeting of expert review should be assessed prospectively in a real clinical setting, where its impact on expert time use and error detection can be measured. Furthermore, the rapid development of multilingual and medically specialised LLMs offers an opportunity to assess whether specialisation improves performance, particularly for rarer clinical extraction targets and for smaller language regions.

## Data Availability

The clinical text data underlying this study cannot be shared publicly due to the Finnish Act on the Secondary Use of Health and Social Data (552/2019) and HUS datagovernance policies; requests for access to the data should be directed to HUS or Findata. The analysis code, prompt template, and JSON output schema used in this study are available from the corresponding author on reasonable request.

## Acknowledgements

We thank pediatric neurologists Mia Westerholm-Ormio and Pirjo Isohanni and pediatrician Matti Hero for their contribution to curating the pediatric stroke cohort data, and pediatric neurologist Maija Seppä-Moilanen for curating the neonatal stroke cohort data used in this study.

## Notes

**Conflicts of interest.** J.L., J.K., and S.K. are employees of Bayer Oy. S.P. and M.K. are affiliated with Helsinki University Hospital (HUS). The authors declare no other conflicts of interest related to this work.

### Competing Interest Statement

J.L., J.K., and S.K. are employees of Bayer Oy. S.P. and M.K. are affiliated with Helsinki University Hospital (HUS). The authors declare no other conflicts of interest related to this work.

### Author Declarations

This was a retrospective registry study using pseudonymised electronic health record data and was conducted under the Finnish Act on the Secondary Use of Health and Social Data (Act 552/2019), which governs research re-use of health data in Finland and provides a legal basis that does not require individual patient consent for retrospective register research of this type. The study was approved by Helsinki University Hospital (HUS approval HUS/265/2023). This approval covers the use of data from patients under 18 years of age. All analyses, including all LLM inference, were performed within the secure HUS Acamedic computing environment.

